# LncRNA antigens- a novel resource to improve immunotherapy efficacy predictions in Melanoma

**DOI:** 10.1101/2023.05.30.23290735

**Authors:** Sumaira Malik, Aaron Golden

## Abstract

**Background:** ICI (immune checkpoint inhibitor) therapy is one of the most promising treatments for melanoma. ICI response however varies among patients, emphasizing the importance of identifying genomic biomarkers to predict likely therapeutic efficacy in advance of treatment. We hypothesised that a lncRNA based immunogencity (lnc-IM) score could be used to predict individual response to ICI treatment, and that this could complement the existing criterion for ICI selection based on tumor mutation burden (TMB).

**Methodology:** The TCGA-SKCM (n=101) and the ICI treated UCLA (n=25), MSKCC (n=16) and DFCI (n=40) melanoma cohorts were used in this study, involving both clinical and transcriptomic data. Each patient was assigned an lnc-IM score based on the number of lncRNA sORF derived peptides predicted to be presented by their tumor’s MHC-I genotype. For the ICI treated cohorts, a combined antigen score was defined as a sum of neo-antigen load (derived from TMB) and lnc-IM score. A logistic regression-based classifier was used to predict ICI responses based on these combined antigen scores.

**Results:** Survival analysis showed improved overall survival among patients with low lnc-IM scores (HR= 0.39, p=0.009) in the TCGA-SKCM cohort. We also observed a negative association between tumor immune cell concentration and lnc-IM scores, with low lnc-IM groups showing higher anti-tumor immune cell concentrations . Using the ICI treated cohorts, we demonstrated that a classifier based on combined antigen scoring improved the prediction of immunotherapy outcomes as compared to using TMB alone, yielding an area under the curve (AUC) of 0.71 with an accuracy of 0.54 and recall of 1. We also demonstrated a reduced rate of false negatives (14%) by using a combined antigen score as compared to the use of TMB alone (33%) in ICI treated cohorts.

**Conclusion:** Our findings suggest that the use of combined antigen scores (using lnc-IM scores along with TMB derived neoantigen load) have potential in improving immunotherapy efficacy predictions. Prospective validation in larger cohort sizes is warranted.

**KEY MESSAGES:** *What is already known on this topic:* Previous studies have established actionable associations between TMB neoantigen load and immunotherapy responses.

*What this study adds:* This study introduces lnc-IM scores as a novel metric that predicts patients antigen load based on translatable lncRNAs expression. These lnc-IM scores when combined with TMB associated neoantigen load indicate an improvement in immunotherapy efficacy predictions.

*How this study might affect research, practice or policy:* Future research is needed to further validate lnc-IM scores as a predictive biomarker for immunotherapy response in various cancer types. The use of lnc-IM scores can empower clinicians to make more informed decisions about administering immunotherapy treatments, improvingpatient outcomes.

## INTRODUCTION

Self versus non-self discrimination by T cells is a hallmark of cancer evasion by the immune response [1]. T cells eliminate cancer cells by recognizing tumor-specific antigens presented on the tumor cell’s surface by MHC-I molecules [2]. Immune and tumor cells possess “checkpoint proteins” such as PD-1, CTLA4 and PDL1 that keep such immune responses in check with the binding of such checkpoint proteins restricting T cells from killing tumor cells. Immune checkpoint inhibitor therapy (ICI) is a promising immunotherapy that restores the T cells’ capacity to attack and eliminate tumor cells by blocking such checkpoint proteins [3].

The use of ICI therapy has been a significant achievement in the last decade for cancer treatment and has demonstrated clear improvements in the survival rate of cancer patients. Its use to date has been approved for multiple cancer types, including melanoma - one of the first cancers to be treated with this therapy [4]. Despite some remarkable successes, response to ICI therapy varies widely among individual patients, emphasizing the importance of identifying genomic biomarkers to try and predict an individual’s likely response in advance of treatment. Tumor mutation burden (TMB), programmed cell death ligand 1 (PDL1), and mismatch repair defect (dMMR)/microsatellites Instability (MSI) are some of the currently FDA-approved predictive biomarkers of ICI efficacy, of which TMB is the most widely used [5,6]. High TMB has been shown to predict improved therapeutic efficacy to ICI therapy in various cancer types [7]. High TMB is associated with a high neoantigen burden [7]; hence, ICI therapy for patients whose cancers have a high TMB would be expected to elicit significant T cell responses against those antigens, resulting in enhanced tumor cell attrition. Although many trials have demonstrated the usability of TMB in clinical practices, a recent study has questioned the concept of the universal usage of TMB as a predictor of ICI efficacy [8], arguing that TMB prediction power is only accurate to a subset of patients, and using TMB as a sole predictor of ICI response might deprive potential patients who might otherwise respond to ICI therapy .

TMB-associated antigens (neoantigens) are derived from non-synonymous missense mutations and originate from the coding region of the genome that encodes proteins [9]. However, TMB is not the only source of antigens. The Encyclopedia of DNA Elements (ENCODE) revealed that the human genome contains almost 80% of non-coding RNAs in contrast to the 1.5% that encodes for proteins [10]. Among non- coding RNAs, those longer than 200 nucleotides in length - collectively known as long non-coding RNAs or lncRNAs - are of great interest on account of their evident wide functional diversity [11]. LncRNAs have been studied for their ability to control regulatory and cellular processes. LncRNAs regulate gene expression both as miRNA sponges and mRNA sponges [12]. LncRNAs act as transcription regulators by modifying the chromatin complexes and can activate or repress gene expression. Moreover, they also control the binding of transcription regulatory factors leading to the activation of nearby genes [13].

LncRNAs have also been shown to be implicated in the creation of MHC-I associated peptides with additional work demonstrating that a considerable number of tumor-specific antigens originate from non-coding regions of the genome [14]. LncRNAs’ contribution to the cancer immunopeptidome is a topic of active research with a particular focus being an assessment of whether such lncRNA-associated antigens are associated with elevated cytotoxic T lymphocyte (CTL) responses. High CTL responses have been associated with better survival and treatment outcomes [15]. Many lncRNAs possess intact short open reading frames (sORF) that can result in the translation of short peptides in the dysregulated cancer transcriptome. Whilst short peptides have been detected in mass-spectrometry based proteomic studies [16], direct association of lncRNA sORFs with tumor immune microenvironment (TIM) has yet to be fully characterised.

In this study we explore how the expression of lncRNA sORFs is associated with a patient’s likely response to ICI therapy. Specifically, we derive a novel metric, the lnc-IM score, that estimates the level of sORF derived peptide presentation based on a tumour’s specific MHC-I genotype. In the first part of our study, we establish the association of lnc-IM scores with the tumor immune microenvironment – determined from known cellular biomarkers - and survival predictions in a melanoma cohort (TCGA-SKCM) without any ICI treatment. In the second part, we utilize three ICI-treated melanoma cohorts to examine how these lnc-IM scores can be used to improve ICI efficacy predictions by their integration with each patient’s TMB-associated antigen count.

## METHODS

### Data acquisition

For this study, three different publicly available melanoma cohorts were utilized. Clinical information and transcriptomic profiles (RNA-seq) of the TCGA-SKCM cohort were downloaded from The Cancer Genome Atlas (TCGA, https://portal.gdc.cancer.gov). HLA typing of the TCGA-SKCM cohort was acquired from a previous study [17]. A total of 101 patients were included in our analysis based on the availability of MHC-I genotype and transcriptomic data and having basic clinical information of age, gender, and overall survival. For ICI efficacy predictions, three ICI-treated cohorts involving metastatic melanoma (UCLA) [18], melanoma (MSKCC) [19] and metastatic melanoma (DFCI) [20] patient groups were utilized. Clinical information, including overall survival, treatment response, TMB, neoantigen load, and transcriptomic data, was retrieved from cBioPortal (https://www.cbioportal.org/) [21]. Based on the criteria mentioned above, a total of 16 patients from the MSKCC cohort, 25 from the UCLA cohort and 40 from DFCI cohort were selected for subsequent analysis. Tumor immune microenvironment (TIM) analysis for the TCGA-SKCM cohort was performed using the xCell algorithm [22], which estimates the the abundance of each immune cell type using expression profiles of specific gene signatures for each cell type.

### Defining Lnc-IM scores

A lncRNA-immunogenicity score (lnc-IM) for a patient was defined as the total number of presentable lncRNA associated sORFs for that patient’s tumor. In the first step, all lncRNAs associated with short open reading frames (sORFs) were retrieved from sORFs.org [23]. We selected our desired dataset from sORFs.org based on filters (*species: humans and biotype = lncRNA*). This initial search resulted in 425 lncRNAs and will be referred to as translatable lncRNAs in this work. These lncRNAs are associated with ∼ 3000 sORFs, as experimentally proven by different Riboseq experiments [23]. Among these translatable lncRNAs, only overexpressed lncRNAs were considered for subsequent analysis. The raw RNA-seq expression of translatable lncRNAs was acquired from GDC (TCGA, https://portal.gdc.cancer.gov) and cBioPortal (https://www.cbioportal.org/). Raw counts were converted to counts per million (cpm) and log normalized using the edgeR [24] package in R. A translatable lncRNA was considered for immunogenicity scoring if it passed the criteria of (log cpm > 6) (Supplemental Figure 1). For each translatable lncRNA, all of its sORFs were considered for lnc-IM scoring. The Patient Harmonic-mean Best Rank (PHBR) score [17] was then assigned to each sORF, which is an estimate of its derived peptide’s MHC-I presentation likelihood. The PHBR score represents the harmonic mean of best-ranked peptides (across a given patient’s HLA alleles). A sORF with PHBR < 0.5 was used to define a “presentable sORF”. The resulting lnc-IM score (Supplemental Figure 2) represents the total number of presentable sORFs in a given patient’s tumor.

### Combined antigen score

For the ICI-treated cohorts, a combined antigen score was derived as a sum of each patient’s TMB associated neoantigen burden (using the methodology previously articulated by [18, 19, 20]) and lnc-IM score (as previously defined by us). Any overlapping loci between sORFs and neoantigen loci were identified and removed to avoid the repetition of potential antigens.

### Statistical Analysis

All analyses were implemented using R (v.3.6.3). Patients were divided into high, low lnc-IM and combined antigen groups using the *surv_cutpoint* and *surv_categorize* functions from the *survminer* package in R [25]. Cutpoints for dividing data into high/low immunogenic groups based on lnc-IM scores and combined antigen scores were chosen for each cohort separately using maximally ranked statistics (Supplemental Figure 2 and Supplemental Table 1). To determine the prognostic value of lnc-IM scores, Kaplan Meier survival plots were generated using the *survival* R package [26]. Spearman’s correlation and Wilcoxon test were conducted using the *ggpubr* [27] R package to determine the association of lnc-IM scores with tumor immune cells. A significance level of 0.05 was used as the cutoff, with p < 0.05 considered as the statistically significant difference for all tests.

Three ICI-treated cohorts were utilized to determine the predictive value of combined antigen score. Patients were stratified into the high and low combined immunogenic groups as described above, and a logistic regression-based classifier was utilized to predict immunotherapy responses using *stats* [28] package in R. Patients were randomly assigned to training 70% and test 30% groups. For performance assessment, the evaluation metrics area under the curve (AUC), accuracy and recall were calculated using R packages *caret* and *caTools* [29, 30]. We compared the prediction power of combined antigen scores with TMB using overall response rates, true positive rates and false negative rates.

## RESULTS

### Lnc-IM scores are associated with anti-tumor immune responses

The tumor immune microenvironment (TIM) is critical in understanding disease progression and predicting treatment responses. Tumor infiltrating lymphocytes (TILs) comprises of a complex set of cells in the TIM that play important roles in both tumor progression and suppression [31]. Based on these roles, these cells can be divided into anti-tumor and pro-tumor immune cell groups. Tumor-associated antigens are known to enhance TILs associated immune responses against tumor cells [31]. To this end, we investigated the association of lncRNA antigen load using lnc- IM scores with abundance of aDC, B cells, macrophages (M1 and M2), CD8-T cells, CD4 memory T-cells, T regulatory cells, Th1 and Th2 cells. Six of these cell types were characterized as anti-tumor immune cells based on their known involvement in tumor surveillance mechanisms, and three were characterized as pro-tumor immune cells. Three of six anti-tumor immune cells showed significant differences between the high and low lnc-IM groups. None of the pro-tumor immune cells showed a significant association between the two groups (Figure 1). Taken together these data show a significant association between lnc-IM scores and elevated anti-tumor immune responses.

**Figure 1:**
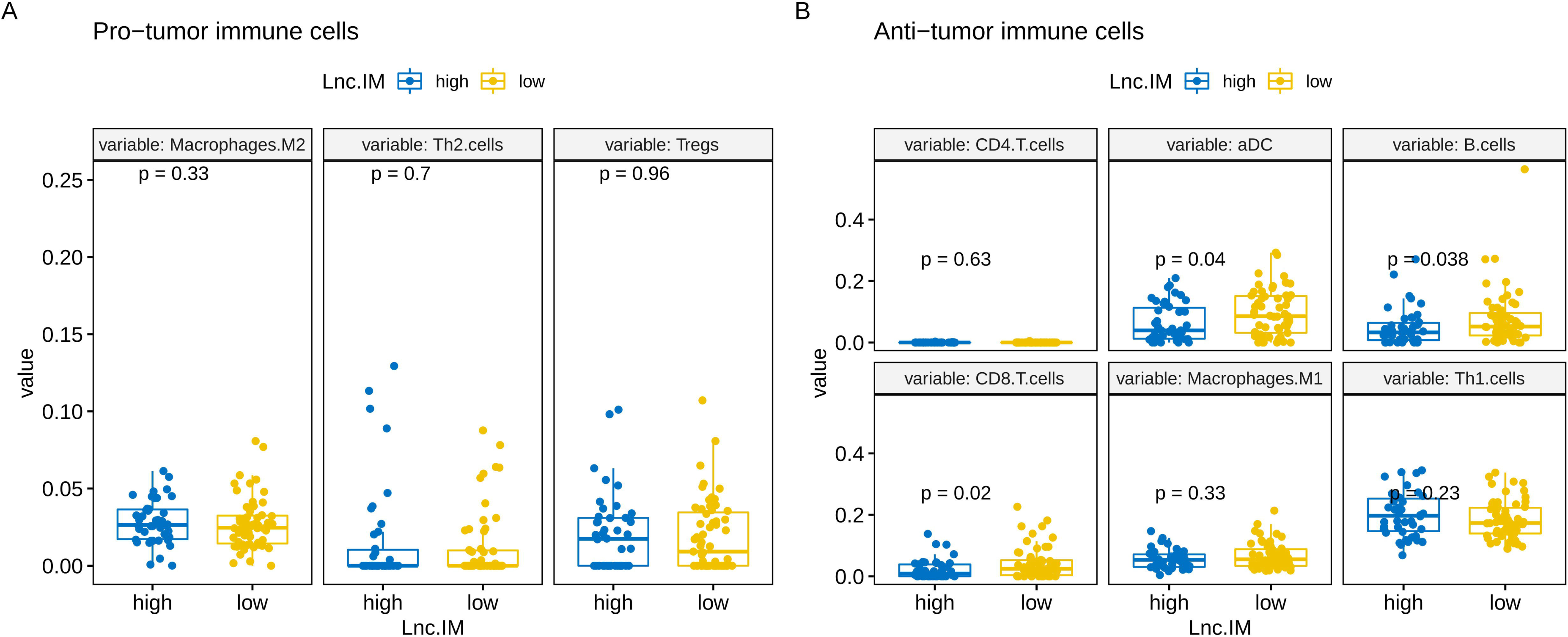
Comparison of immune cells abundance between high and low immunogenic groups. A) Pro-tumor immune cells show no significant association with lnc-IM scores. B) Association of anti-tumor immune cells with high and low lnc-IM scores.

### Lnc-IM scores are associated with survival predictions

High tumor infiltrating lymphocytes (TILs) levels have been associated with the immune system’s capacity to eliminate tumor cells [32]. Hence, patients with high TILs show improved survival compared to those with low TILs. Knowing the value of TILs in survival prediction [33] and the association of lnc-IM scores with TILs (Figure 1), we next questioned if lnc-IM scores could be used to predict survival in the TCGA-SKCM cohort. The results showed that patients in the low lnc-IM category (n = 59) showed better overall survival (HR = 0.39, p = 0.009) than in the high lnc-IM category (n = 42) (Figure 2). The overall median survival remained at 1070 days among the low lnc-IM group, while it reduced to 721 days in the high-IM group. The baseline characteristics of all patients are shown in Table 1. In order to check the association of any confounding variables with survival predictions, a multivariate analysis was performed using age, gender, and cancer stage and lnc-IM scores (Figure 3). None except lnc-IM scores, were significantly associated with survival predictions.

**Figure 2:**
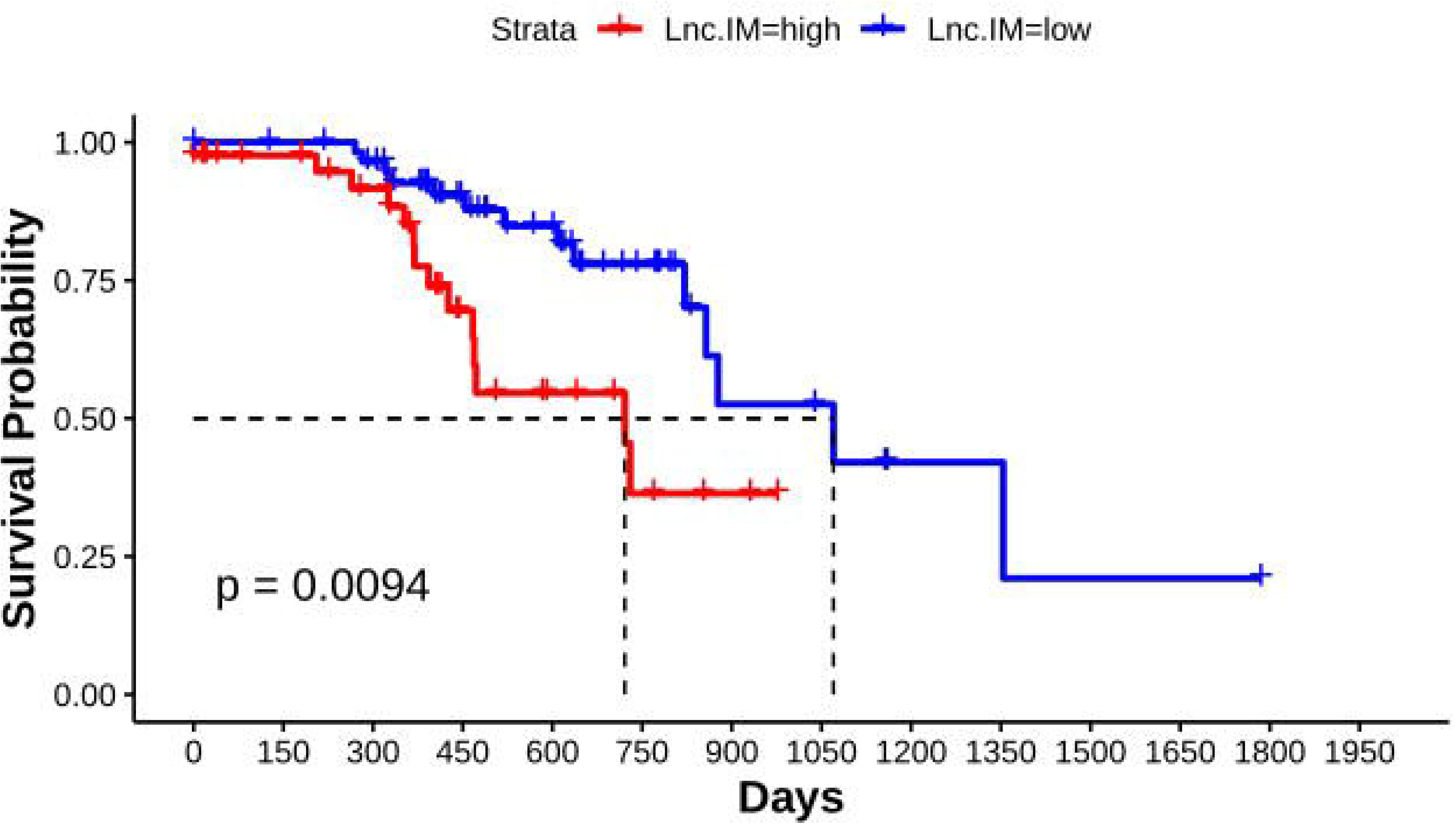
Association of lnc-IM scores with survival predictions in TCGA-SKCM cohort (n=101). Low lnc-IM group (n=59) is associated with better survival outcomes as compared to high lnc-IM group (n=42).

**Figure 3:**
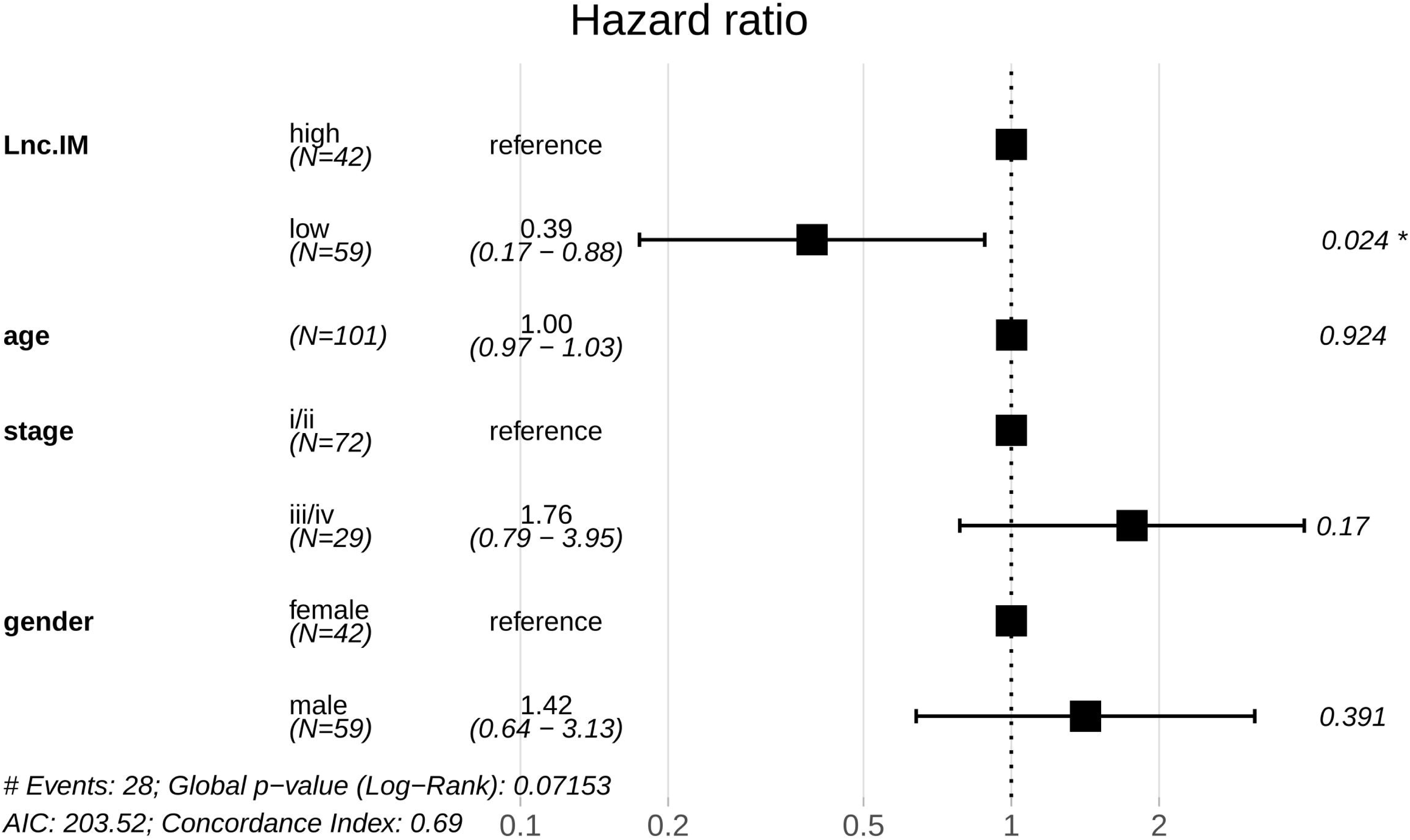
Multivariate analysis to identify any confounding variables. Only immunity count (lnc-IM scores) showed significance with survival predictions (TCGA-SKCM).

**Table 1:** Baseline features of cohorts used in this study. For ICI treated cohorts MSKCC, UCLA only subset of patients was selected for downstream analysis (as explained in Methods: Data acquisition).

|  | TCGA-SKCM | UCLA | MSKCC | DFCI |
| --- | --- | --- | --- | --- |
| **Gender** |  |  |  |  |
| **Female** | 42 | 11 | 25 | 14 |
| **Male** | 59 | 27 | 39 | 26 |
| **Age** |  |  |  |  |
| **≤50** | 15 | 7 | 14 | 12 |
| **>50** | 86 | 31 | 47 | 28 |
| **Treatment** |  |  |  |  |
| **Iplimumab** | - | - | 60 | 40 |
| **Pembrolizumab** | - | 36 | - | - |
| **Nivolumab** | - | 2 | - | - |
| **Tremelimumab** | - | - | 4 | - |
| **Responders** | - | 21 | 14 | 5 |
| **Cancer stage** |  |  |  |  |
| **I/II** | 72 | - | - | - |
| **III/IV** | 29 | - | - | - |

### ICI efficacy predictions based on combined antigen score

Based on the association of lnc-IM scores with both TILs and survival, we evaluated how such scoring can help improve immunotherapy outcomes prediction. We hypothesized that using a combined antigen score that includes both lncRNA- associated antigen scores and TMB-associated neoantigen scores can give a fuller picture of the tumor’s immunopeptidome and so enhance the prediction of ICI efficacy. Such scoring could also help predict ICI outcomes for patients’ whose tumors are not hypermutated (i.e have a high TMB) and so would not be typically considered for such treatment. Using both lnc-IM scores and TMB-derived neoantigen load, a combined antigen score was assigned to each patient in the ICI-treated cohorts. A logistic regression-based classifier was then adopted to build a prediction model for immunotherapy outcomes in a combined cohort.

We evaluated the model’s performance; our results showed an overall AUC of 0.71 in discriminating between responders and non-responders (accuracy = 0.54 and recall = 1). We compared this model with TMB based model (FDA approved cutoff > 10 mutations/megabase (mut/Mb) was used for dividing patients into high and low TMB groups). The results showed an improved performance of combined antigen scoring as compared to TMB based model (Table 2).

**Table 2:**
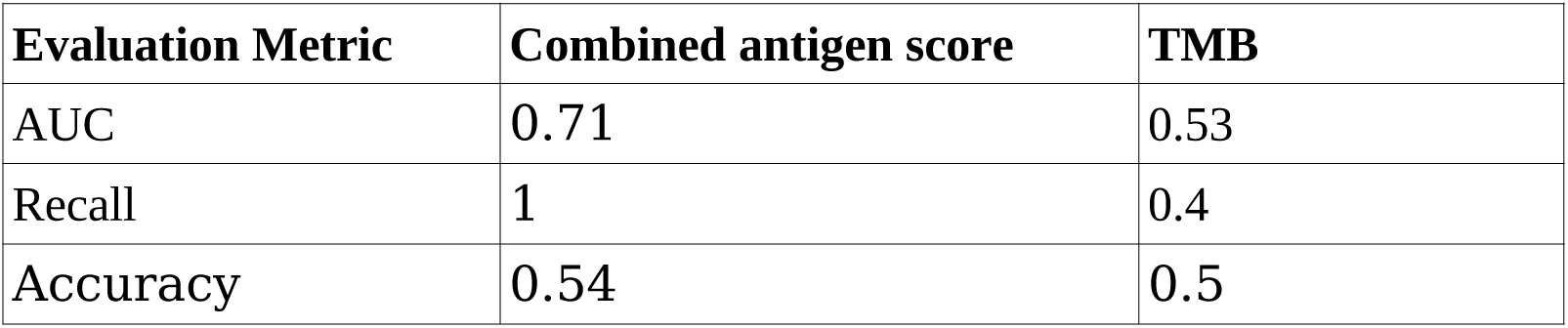
Evaluation metrics for combined antigen scoring and TMB based models.

### Combined antigen score performs better than TMB alone as biomarker

To further assess whether predictions based on combined antigen score performed better than using TMB alone, we compared overall response rates (ORR) between the high TMB group and high combined antigen score in all three ICI treated cohorts. The ORR improved in high combined antigen score as compared to high TMB among UCLA and MSKCC cohorts while remaining the same for DFCI cohort (Figure 4). Among these ICI treated cohorts, 21 out of 81 patients responded to therapy. Using a high combined antigen score (cutoff criterion showed in Supplemental Material Table 1) as a classifier, 18 out of these 21 were correctly identified as responders. In contrast, using TMB alone using the same classifier formalism correctly categorizes only 14 responders. An additional advantage of using a combined strategy is the goal of minimizing the false negative rate to ensure a robust classification that would not deprive potential responders of therapy. In Figure 5 we show that the false negative rate decreased from 33% to 14% using combined antigen score than TMB alone. These results demonstrate strong evidence that using combined antigen score can help improve prediction efficacy for patients who might have a low mutation burden but still can benefit from treatment.

**Figure 4:**
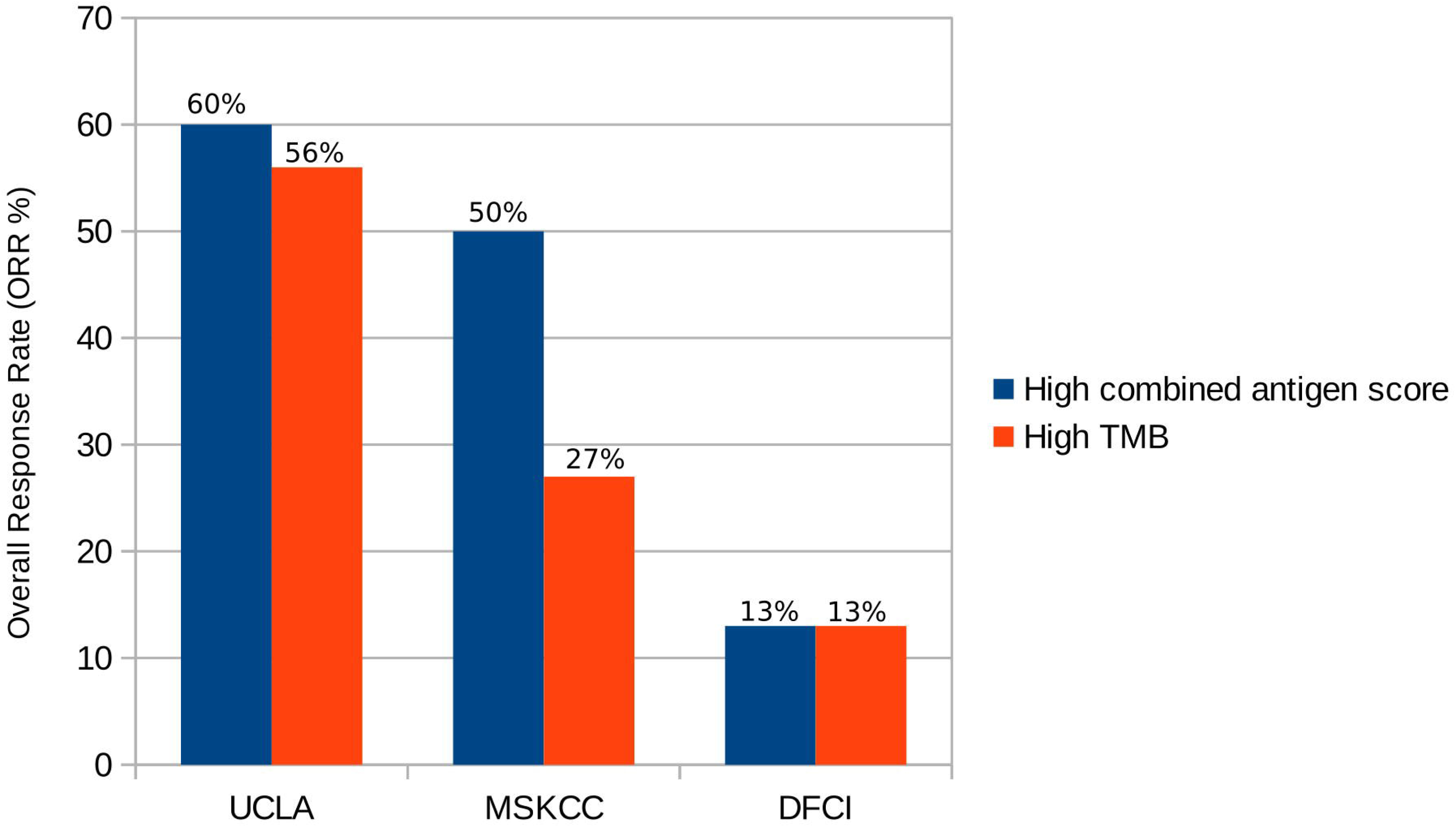
Comparison of overall response rate between high TMB category and high combined antigen score among all ICI treated cohorts.

**Figure 5:**
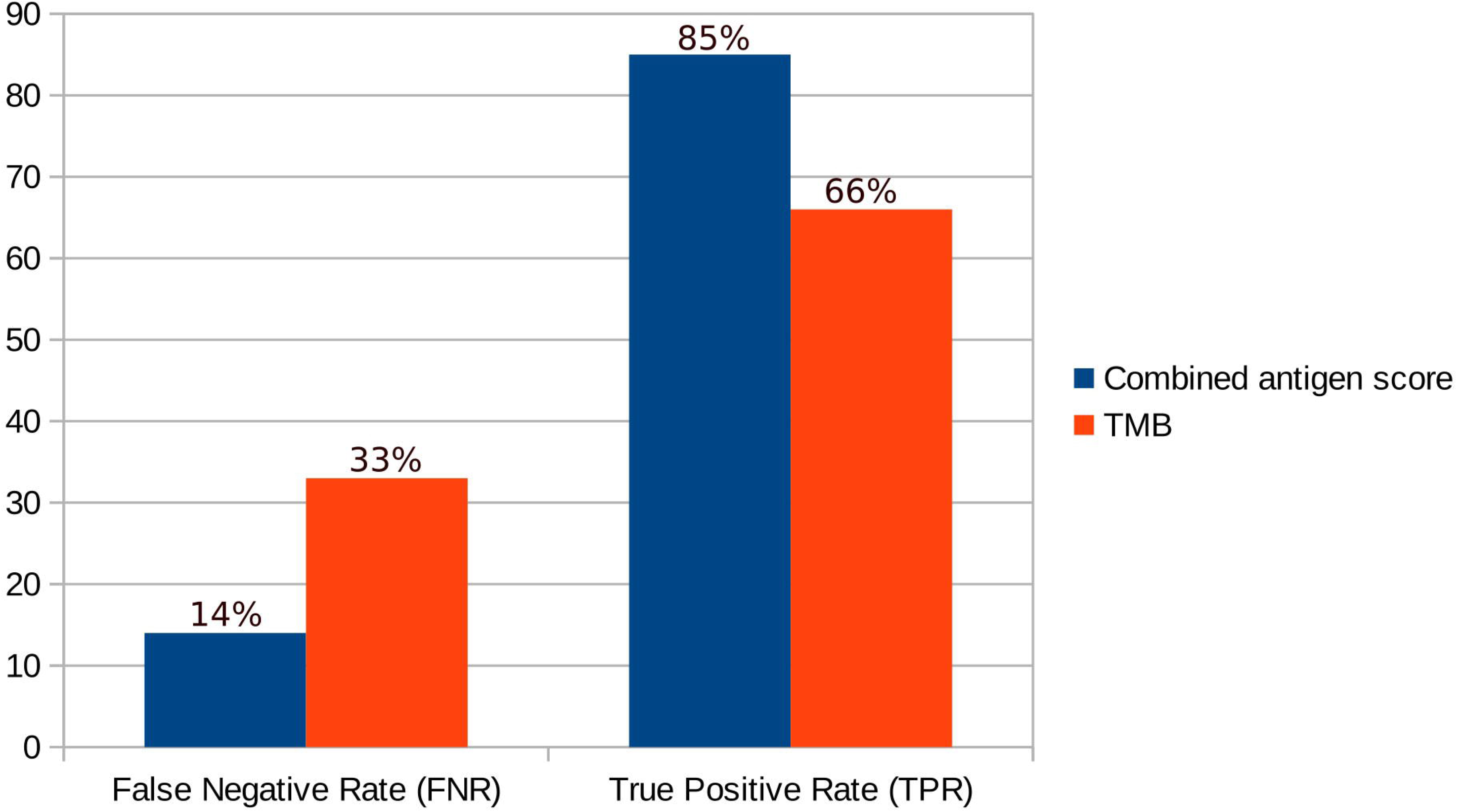
Comparing true positive rate (TPR) and false negative rate (FNR) between high TMB and high combined antigen scores. High combined antigen scoring showed improved TPR of 85% as compared to high TMB (66%). FNR also showed improvement (14%) in high combined antigen classifier.

## DISCUSSION

In this bioinformatics-based study, we introduced a novel metric “lnc-IM scores” incorporating translatable lncRNA expression and patient MHC-I genotype and combined them with TMB-associated neoantigen load to create a predictive biomarker “combined antigen score” for ICI outcomes. TCGA SKCM data revealed the prognostic value and association of lnc-IM scores with the tumor immune microenvironment. Additionally, the value of combined antigen score as a predictive biomarker was investigated using ICI-treated cohorts (UCLA n=27, MSKCC n=21, DFCI n=40), previously used to investigate TMB-associated ICI response alone.

Among the TCGA SKCM cohort, lncRNA immunogenicity scores were significantly associated with anti-tumor immune responses. TILs associated with anti-tumor immune responses showed significant differences between high and low lnc-IM groups. Interestingly, all of these cells were upregulated in the low lnc-IM group. These findings appear to be in accordance with recent pan-cancer studies where high TMB/high antigen groups were associated with depressed immune cell infiltration in different cancer types [34]. One of the possible explanations for such trends could be that the quality/immunogenicity of antigen being presented is responsible for a better immune response than the quantity of antigens [35]. A study previously showed that certain antigens belonging to ERVs were associated with high antigen specific CD8-Tcell infiltrate as compared to other antigens that were associated with rare antigen specific CD8-T cells population [15]. Other critical factors are the tumor immune evasion mechanisms that are prevalent in high immunogenic groups. Cancers tend to evade immune responses by downregulating MHC molecules and TAP3 proteins, so these antigens are not efficiently presented on cancer cell surfaces [36]. Hence, we see a decrease in immune cell infiltration in such cancers.

TILs have been linked to prognostic outcomes in various types of cancer. In general, elevated levels of TILs within a tumor correspond to a more aggressive anti-tumor immune microenvironment and are associated with better survival outcomes compared to tumors with low TILs, as previously reported [37]. Our study (as illustrated in Figure 1 and Figure 2) supports this finding by demonstrating that patients with low lnc-IM tumors generally had greater anti-tumor TILs and better survival outcomes than those with high lnc-IM tumors.

We hypothesized that using lnc-IM scores along with conventional TMB might help improve efficacy predictions. We determined that the overall response rates improved using high combined antigen score as compared to using TMB alone. An essential factor is to ensure that the combined antigen score based classifier helps decrease the false negative rate to avoid depriving actual responders of ICI therapy. We have demonstrated that using the high combined antigen score compared to high TMB alone cannot only improve the true positive rate but also help decrease the false negative rate (Figure 5). In these cohorts, using high TMB alone as a biomarker could deprive 33% of potential responders of potentially efficacious ICI therapy. However, in our small study we demonstrated by using high combined antigen scores reduced this percentage to 14%. These findings have specifically highlighted the predictive power of combined antigen approach scores for responders with low TMB but high lnc-IM scores.

The key limitations of this study are the small sample size, the retrospective nature of cohorts used, and the variability of the type of ICI treatment administered. ICI treatments vary based on the type of checkpoint protein targeted (PD-1 or CTLA4). Both treatments show potential for melanoma treatment with 58% ORR for anti-PD-1 and 38% ORR for anti-CTLA4 therapy in melanoma [38]. Using both primary (TCGA-SKCM) and metastatic cohorts (ICI treated) in our study may have advantages and limitations. On the one hand, the inclusion of both sample types increased the sample size of the study, enhancing our ability to identify more inclusive and robust prognostic and predictive biomarkers. On the other hand, using both cohorts may present challenges due to the potential differences in molecular profiles between primary and metastatic tumors. We note however, a recent study showing no significant difference between primary and metastatic melanoma TMB [39]. We have demonstrated an association of the lnc-IM score with the TIM, survival, and ICI outcomes prediction in three different cohorts providing confidence as regards the added value of the lnc-IM scores as a biomarker. It is also worth noting that the predictive power of combined antigen scoring was evaluated specifically in metastatic tumors. Further validation in a larger cohort size is required to determine the optimal cutoff for the lnc-IM and combined antigen scores.

Future research would ideally focus on determining the value of lnc-IM scores in other cancer types that are not hypermutated yet still respond to ICI therapy, involving larger cohort sizes.

## Data Availability

All data produced in the present study are available upon reasonable request to the authors

https://portal.gdc.cancer.gov

https://www.cbioportal.org/

## Acknowledgements

This project was supported by a Hardiman Research Scholarship, University of Galway, Ireland

## Ethics approval

All the data used in this study were derived from existing publicly available databases and previously published studies. All the data were pre-anonymized. Thus, ethical approval was not required.

## Data availability statement

Data can be provided on request.

## List of Abbreviations

ICI: Immune checkpoint inhibitor therapy
lnc-IM score: LncRNA based immunogencity score
TMB: Tumor mutation burden
AUC: Area under the curve
PDL1: Programmed cell death ligand 1
dMMR: Mismatch repair defect
MSI: Microsatellites Instability
ENCODE: Encyclopedia of DNA Elements
CTL: Cytotoxic T lymphocyte
sORFs: Short open reading frames
TIM: Tumor immune microenvironment
TCGA: The Cancer Genome Atlas
cpm: Counts per million
PHBR: Patient Harmonic mean Best Ran
TILs: Tumor infiltrating lymphocytes
ORR: Overall response rates
HR: Hazard ratio

