## Supplemental Figure 1 for "LncRNA antigens- a novel resource to improve immunotherapy efficacy predictions in Melanoma"

**Supplemental Material**

**Supplemental Figure 1: Cutoff selection for lncRNA expression**

In order to ensure only overexpressed lncRNAs are chosen for the subsequent formulation of the lnc-IM scores, lncRNAs with logcpm > 6 were chosen. As we can see in this figure, average expression is marked around logcpm 5; chosen threshold will ensure lowly or average expressed lncRNAs are filtered.

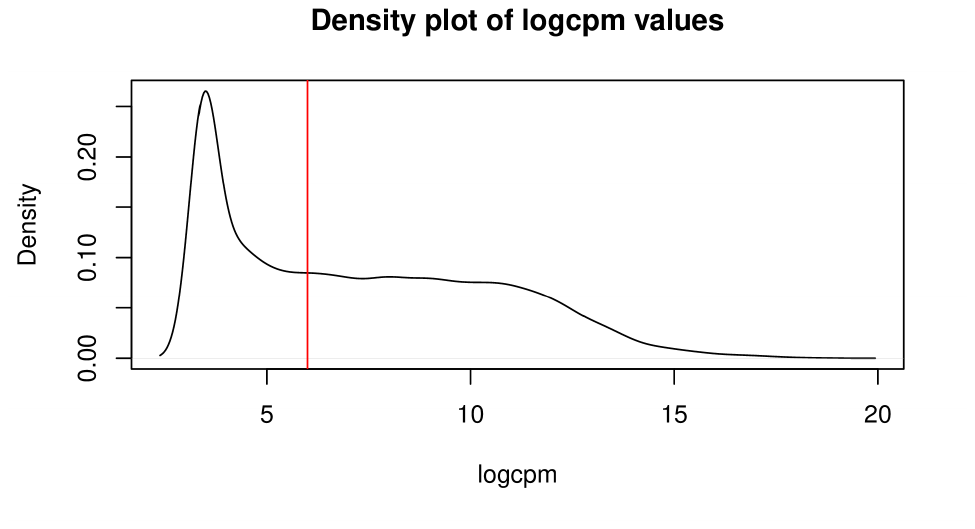

Figur Figure 1: Cutoff selection for overexpressed translatable lncRNAs. The cutoff logcpm > 6 was selected for subsequent analysis.

**Supplemental Figure 2: Cutoff selection for high and low lnc-IM groups (TCGA-SKCM)**

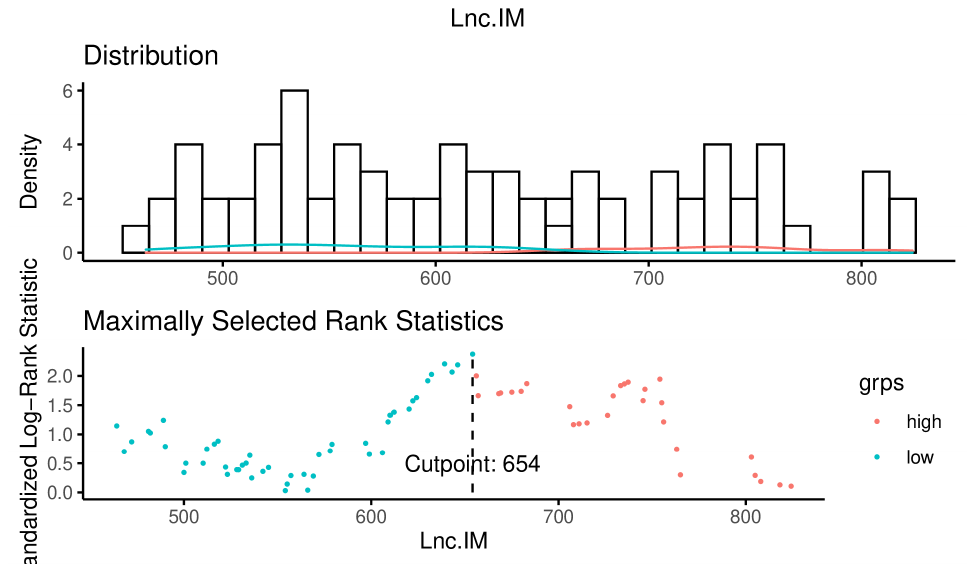

Figure 2: We used maximally selected rank statistic approach to identify appropiate cutoff to divide patients into low and high lnc-IM groups. Lnc-IM scores range from 320-990 and patients with lnc-IM counts > 654 were characterized as high lnc-IM group. Total 59 patients were characterized in low lnc-IM and 42 in high lnc-IM groups.

**Supplemental Table 1: Selected cutoff for combined antigen count in ICI treated cohorts defined by maximally selected rank statistic**

| **Cohort** | **Data range** | **Cutoff** |
| --- | --- | --- |
| **UCLA** | 153-5138 | 389 |
| **MSKCC** | 515-1697 | 1252 |
| **DFCI** | 250-4130 | 364 |
